# Incidence and Epidemiological study of COVID-19 in Nagpur urban region (India) using Molecular testing

**DOI:** 10.1101/2021.05.11.21256719

**Authors:** Jay Tanna, Bishwadeep Singha, Amit R. Nayak, Aliabbas A. Husain, Dhananjay V. Raje, Shubhangi A. Desai, Madhavi Deshmukh, Shailendra Mundhada, Rajpal S Kashyap

## Abstract

The COVID-19 pandemic caused by severe acute respiratory syndrome coronavirus-2 (SARS CoV-2) virus has emerged as public health emergency affecting 206 countries worldwide. India is second highest currently worst effected by Covid 19 pandemic with close to 12.6 million cases and 1.6K deaths reported till date. Maharahstra is the highest Covid-19 burden state in India reporting quarter of overall cases. The city of Nagpur, in Maharashtra state, ranks 4th in terms of reported COVID-19 cases, with 2.5 lakh incidences and more than 4,000 deaths As the transmission rate of COVID-19 is high, it is imperative to study its disease epidemiology in regions of high endemicity to bolster our understanding of its spread, transmission dynamics and contact tracing to undertake appropriate public health control measures.. The present study was undertaken to study the incidence and trend of COVID-19 infection from various zonal regions of Nagpur city, using real time PCR (RT PCR). A retrospective study was carried out at Indian Council of Medical Research (ICMR) approved private molecular diagnostic laboratory in Nagpur from period of 4th May 2020 to 14th November 2020. A total of 51,532 samples collected from various zonal regions of the city during the study period were processed for SARS CoV-2 RT-PCR. Patient information was collected using a pre-defined study proforma which included demographic details such as name, age, gender, address, along with other information, like details of sample collected, kits used and date of sample collected and processed. The study reports an overall Covid-19 positivity of 34% in Nagpur region. The zone wise distribution of positive cases indicated high rate of COVID-19 in endemic regions of Nagpur such as Satranjipura (49%), Ashi nagar (44%), Gandhibagh (43%) & Lakadganj (43%). Rates of infection were high in economically productive age group (21-40) with males being more vulnerable than females. The result of present epidemiology study highlights important data with respect to regions of endemicity within Nagpur city zones. The present data has high public health importance and will be useful for local civic bodies and other community stake holders to undertake appropriate control measures in future epidemic waves of Covid 19. Interestingly, the Government’s reduction in testing rates has been helpful in increasing testing per day. The authorization of private laboratories has also increased testing.

## Introduction

Severe acute respiratory syndrome coronavirus-2 (SARS CoV-2) is a RNA virus that has caused the COVID-19 pandemic [1, 2]. A total of 132 million cases have been reported worldwide with 2.86 million deaths. India is second highest currently worst effected by Covid 19 pandemic with close to 12.6 million cases and 1.6K deaths reported till date [3.4]. Maharahstra is the highest Covid 19 burden state in India reporting quarter of overall cases [5, 6]. Detection, assessment and rapid response are critical for epidemics and pandemics. In the early stages of the spread, the public health system failed to provide an adequate number of personal protective equipment, testing, contact tracing and shortages of medical equipment. However, successful engagement of public private partnership has been in such a crisis situation [7, 8].

Nagpur is a tier 2 city and is one of the rapidly growing cities of Maharashtra and Central India. It is located at the eastern tip of Maharashtra and is located in the heart of India. Nagpur Municipal Corporation (NMC) is the major municipal body that administers the Nagpur region. NMC split Nagpur into 10 zones, which are also divided into 38 wards [9, 10]. In Nagpur, the first COVID-19 case was reported on March 12, 2020, a 45-year-old male who travelled from the United States. Till December 2020, the city had recorded a total of 98 thousand cases with 2600 deaths. As of March 2021, the city had recorded a total of 2.3 lakh cases [11]

To date, COVID-19 epidemiology has been investigated at the National, and international levels. A micro analysis of Urban Nagpur as an individual tier-2 city is presented and analyzed in this study. Positive cases were segregated in terms of geographic area with postal pin codes within the city jurisdiction. The data are analyzed taking into account the initiatives of the Nagpur Municipal Corporation, the State of Maharashtra and the national government to combat the disease [12]

Understanding the epidemiology of diseases makes it possible to prevent the spread of diseases. Unfortunately, India does not pay much attention to this aspect. The epidemiological study also helps in assessing risk factors associated with the disease that represent critical control points. These studies are important in implementing good public policy decisions. Since the pandemic began, a large amount of data has been generated daily around the world and can be easily assessed by the public. Using these data authorities can structure beneficial policies that can be useful in strengthening the public health infrastructure [7, 8, 13, 14].

The analysis of test data with respect to all of the above parameters is conducted to assess the effectiveness of the various policies. The data and their analysis could help assess the effectiveness of various governmental strategies at the local and national levels to control the spread of the disease. It will also facilitate the development of future strategies for new spikes. It may also be helpful to prioritize the population to be immunized if we can extend this study to antibody screening [15]

## Material and Methods

### Study design and participants

This is a retrospective study of the COVID-19 RT-PCR test that was conducted at the Dhruv Pathology and Molecular Diagnostic Lab from May 4, 2020 to November 14, 2020. The Laboratory is prominent stand alone, National Accreditation Board for Testing and Calibration Laboratories (NABL) accredited diagnostic center in Nagpur, Maharashtra (NABL Registration Number MC2701) and was first private laboratory in Central India, approved by ICMR, Govt of India for COVID-19 RT-PCR testing (ICMR Registration Number DPMDLN). The lab successfully participates in ICMR and WHO external quality assurance programs.

### Sample Collection

The ICMR guidelines were followed for sample collection and analysis (F. No. Z.28015/23/2020-EMR, dated: 21^st^ March 2020).Samples were collected either at the collection centre (dedicates sample collection room) or hospitals (for admitted cases) or at homes by the trained collection personnel. Throat and/or nasopharyngeal specimens were obtained using standard techniques. Care was taken to use only dacron and nylon coated swabs in the collection of samples. Cotton swabs were avoided at any cost to avoid PCR inhibition. The swabs were immediately transferred to a prelabelled viral transport medium (VTM) or viral lysis medium (VLM). The VTM/VLM tubes were placed in zipper bags and stored in a secondary container with ice bags to maintain temperatures. Appropriate cold chain was maintained for transportation to testing center. All samples were processed within 12 hours of receipt at the centre by qualified personnel using real-time RT-PCR technology.

Patient information were collected using a pre-defined clinical proforma which include demographic details such as name, age, gender, address, type of sample, type of kit used and date of sample collected and processed. A total of 51,531 cases were tested in the laboratory from May 2020 to November 2020. Out of 51531 cases, 1486 cases were excluded as these samples were from other states, 6728 cases were excluded as these samples were form other districts or cities in Maharashtra and 7485 cases were excluded as location details for these cases were not available, remaining 35830 cases received from Nagpur region were further distributed into 10 zones on the basis of location details provided while collecting sample for COVID testing.

### Nucleic acid extraction and Real time RT-PCR for SARS CoV-2 for E gene and RdRp gene

The swabs collected in VTM/VLM were further subjected to Nucleic Acid Extraction and amplification. SARS-CoV-2 nucleic acid (RNA) has been extracted through various methods, including manual spin column extraction and automated magnetic ball extraction.

#### Nucleic acid extraction: For RNA extraction, a sample volume of 200µl was used followed by lysis and homogenisation to access RNA in cells

RNA has been separated from other cell components by silicon membrane technology or in the spin column. In addition to manual kits, automated extraction machines was also to reduce the extraction time, optimize yield of RNA, increase the amount and quality of isolated RNA, and reduce chances of cross contamination and of course to increase the rate of extraction. The eluted NRAs were kept at a temperature of 2 to 8°C until amplification.

### Real-time PCR for detection of SARS CoV-2

RT-PCR testing was done using PathoDetectTM COVID19 Qualitative PCR kits from Mylab Discovery solutions which targets Envelope gene (E) and RNA-dependent RNA polymerase gene (RdRp) as per manufacturer’s recommendations. E gene was used for screening of SARS-like Corona viruses. RdRp gene or Orf 1 gene were used for specific detection SARS-COV-2. RT-PCR was carried out using Quant Studio 5 (Thermo Fisher, USA) Internal controls and positive controls were put with every cycle to assure precise amplification cycle. Test results were shared with patients using the contact information provided during sample submission or were asked to obtain the results by visiting the test centre. All the test results were also submitted to governing authority of Nagpur and ICMR health portal as per the guidelines proposed by ICMR for National surveillance data

### Statistical analysis

Data for the Ct-Value was represented as Mean ± SD. Categorical variables were presented as frequency and percentages (n; %). Comparability of groups was analyzed by Chi-square test, Student’s t-test, or Mann-Whitney test as appropriate using the statistical software MedCalc statistical software version 10. A p value<0.05 was considered statistically significant

## Results

### Results

For all samples analyzed, patient demographic characteristics were compiled with descriptive statistics. The descriptive analysis includes age distribution, gender ratio (male: female ratio), mean Ct value, and zonal distribution of cases tested.

Overall, during the 28-week period between May 4, 2020 and November 14, 2020, a total of 51,531 samples were received for COVID-19 testing. After applying exclusion criteria, few cases were excluded from the study as they were not from Nagpur region and were either residents from other states or districts or may have traveling history. After the exclusion of these cases, the final samples in the Nagpur region were 43,317 (Figure 1). Figure 2 shows the total number of samples tested and Male-Female distribution over the period of 28 weeks. It was observed that there was a gradual increase in the testing from the month of June 2020 and the center received the highest number of samples for testing in the month of September 2020. There was a steady decline in COVID-19 RT-PCR testing at the end of September as many other institutes and testing centers were approved for COVID-19 testing in Nagpur by ICMR. The number of male patients was greater than the number of female patients among the cases tested during the 28 weeks.

**Figure 1:**
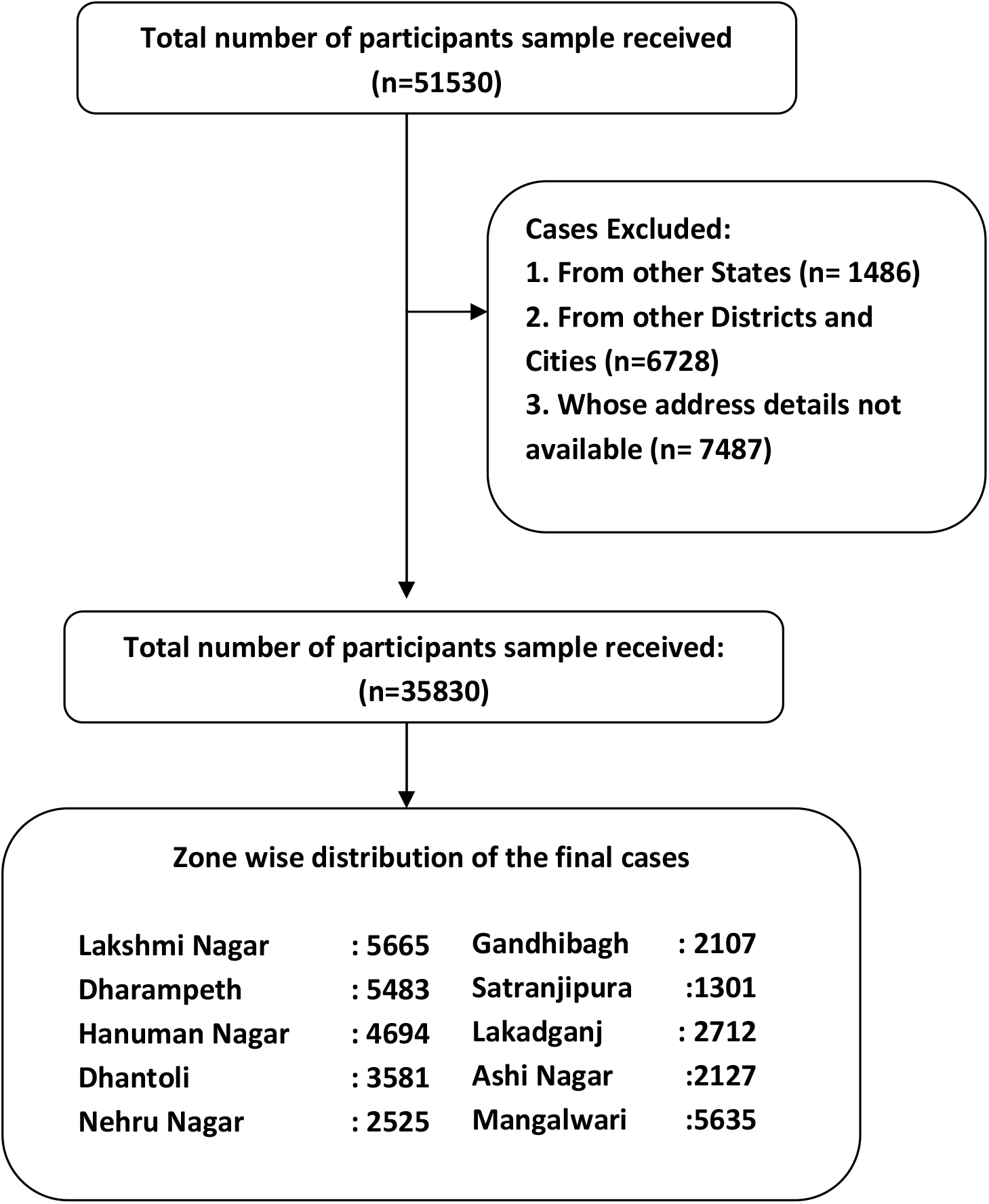
Flow chart for Inclusion and exclusion

**Figure 2:**
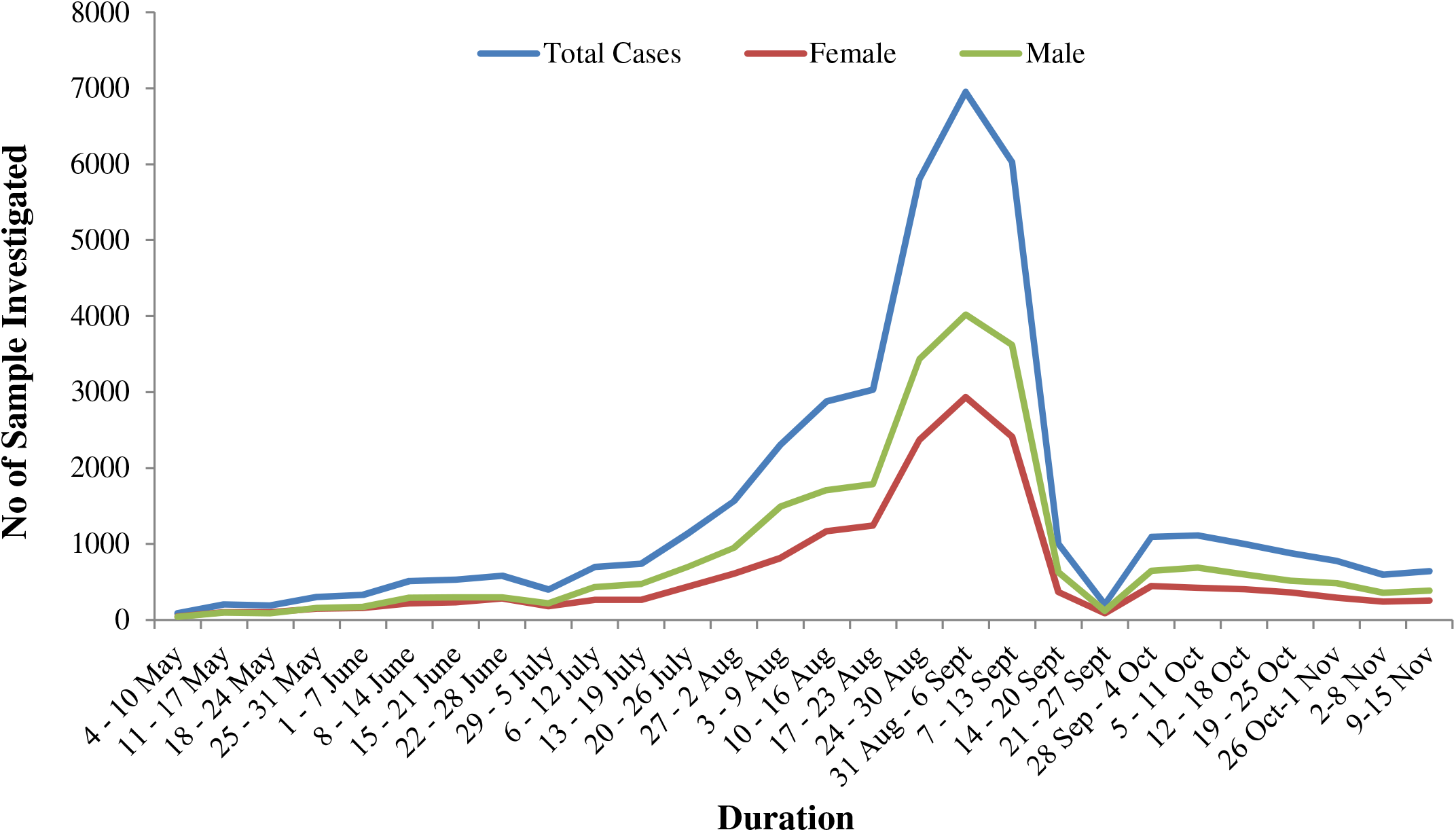
Weekly distribution of total sample tested and Male-Female distribution.

The samples received for testing were divided into age groups of 20 years as shown in Figure 3 and percentage positive cases among these group is shown in Figure 4. The majority of cases occurred in the 21-40 age group (43%) with positivity of 36%, followed by the 41-60 age group (32%) with positivity of 37%. These age groups consist of students and working professionals and are among the most exposed groups. A large number of samples were also received from the age group 61-80 years (13%) with positivity of 16% and 0 to 20 years (10%) with positivity of 10%. Very few samples were received from those older than 80 (2%) with positivity of 1%.

**Figure 3:**
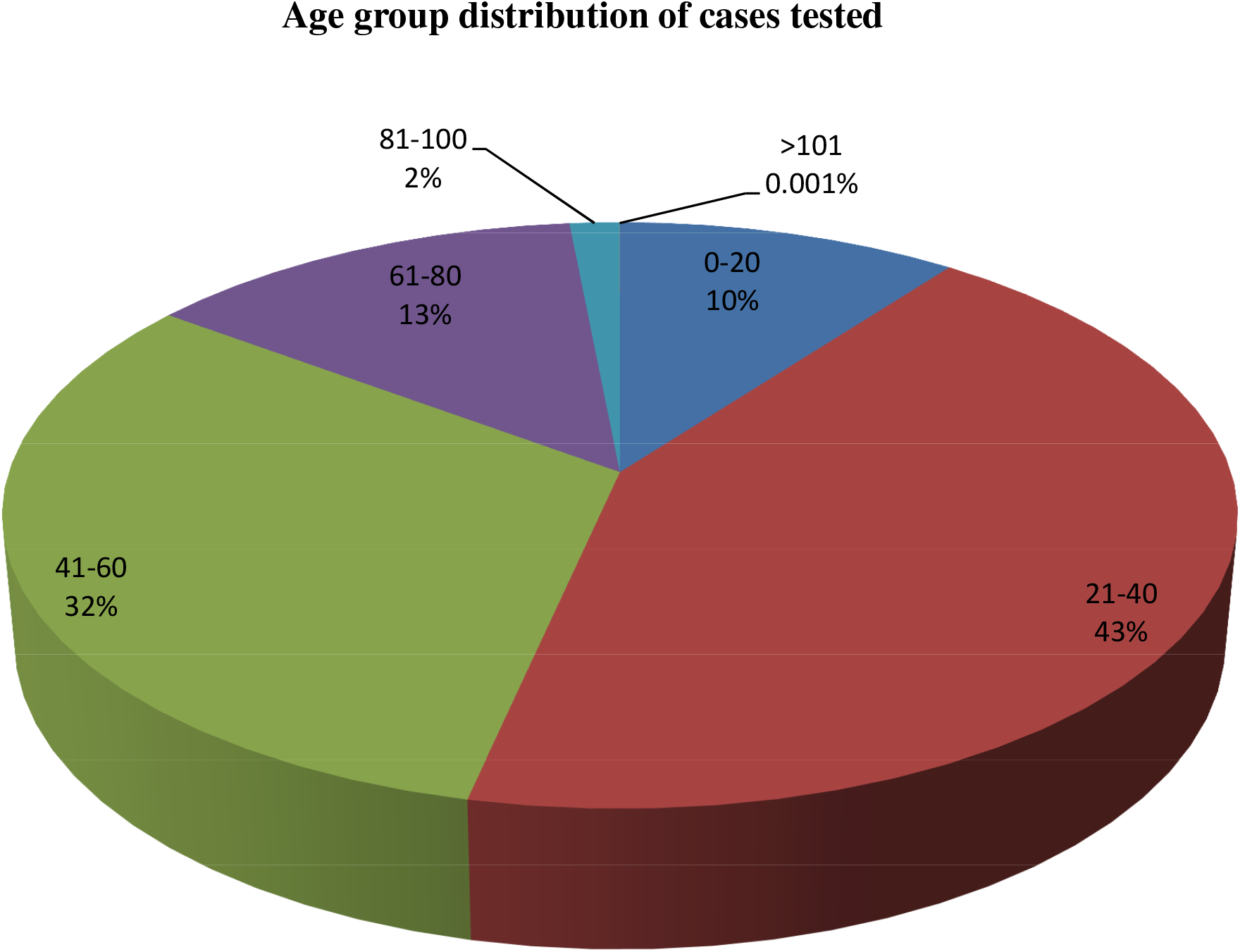
Age group distribution for the cases tested at the Dhruv laboratory for Nagpur region (n=43317)

**Figure 4:**
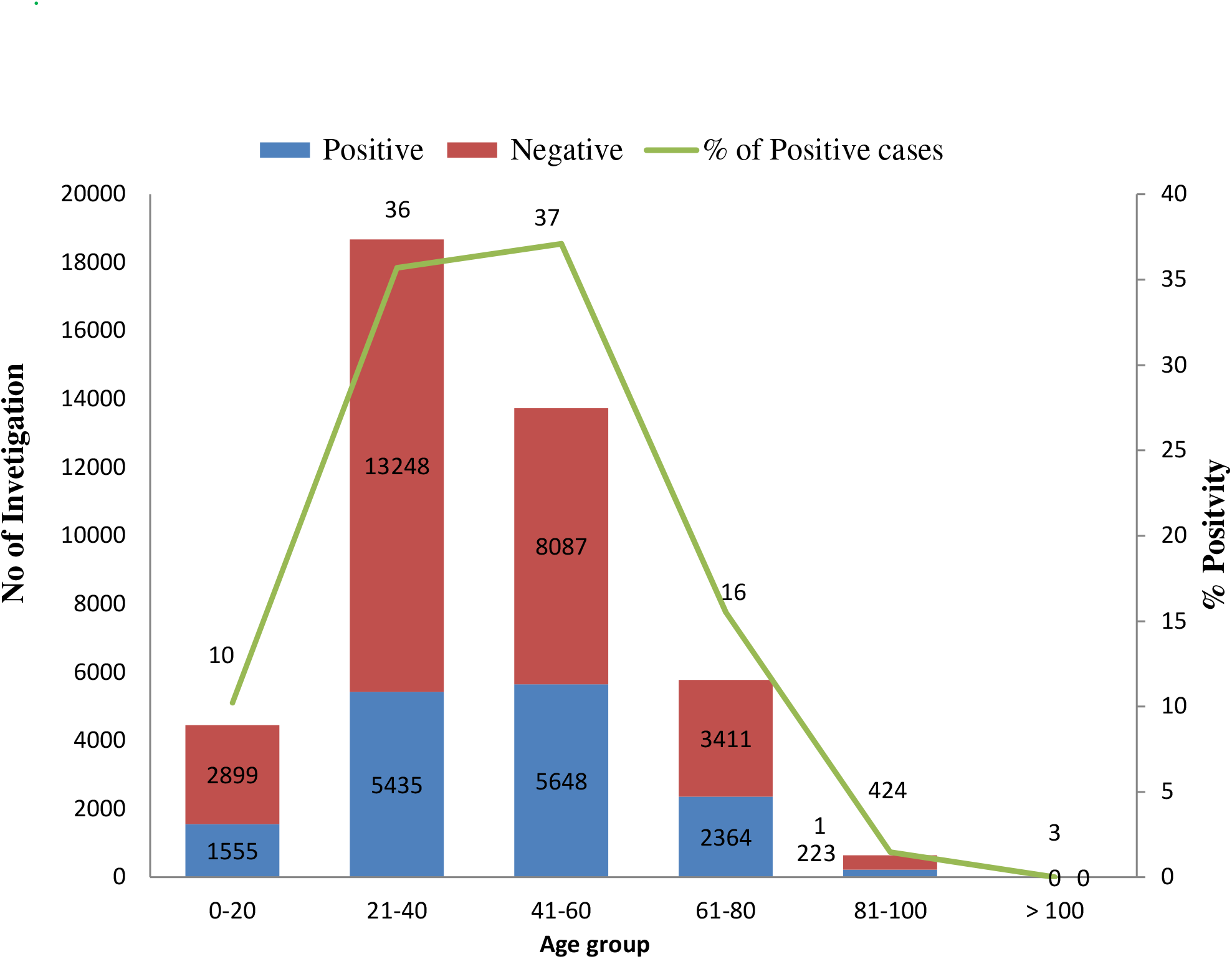
Results of the RT-PCR test among different age group

Results for the cases tested for COVID RT-PCR were categorized into positive, negative and inconclusive on the basis of Ct value cut off. The inconclusive findings were also assessed as negative. Of the 43,317 cases tested, 15,225 were positive (35.15%), 28,072 were negative (64.8%) and 20 were inconclusive (0.05%). The results of the tests were distributed across different age groups, as shown in Figure 4. Significantly high positive cases (P<0.05 vs 0-20Years) were observed for the age group 41-60 years [n=5648 (37%)] followed by age group 21-40 years [n=5435 (36%)], 61-80 years [n=2364 (16%)], as compared to 0-20 years [n=1555 (10%)]. While very less no of cases investigated for age group of 81-100 years [223 (1%)]. Interestingly there was no positive case in age group above 100 years [n=3].

Total samples received from Nagpur urban region was further distributed into 10 zones on the basis of location details provided while collecting samples for COVID-19 testing. Figure 5 shows the percentage distribution of cases tested in different zones.. Result shows that majority of testing (70%) investigated cases were from five zones only [i.e Lakshmi Nagar zone (16%), Mangalwari zone (16%), Dharampeth zone (15%), Hanuman Nagar zone (13%) &, Dhantoli zone (10%)] and 30% investigations were done remaining five zone [i.e Lakadganj zone (8%), Nehru Nagar (7%) zone, Gandhibagh (6%), Ashi Nagar zone (6%) and Satranjipura zone (4%)]. There is similar pattern of a age group of patient investigated from each zone i.e highest cases from age group 21-40 years followed by 41-60 years, 61-80 years, 0-20 years and >80 years (*supplementary Figure S1*). Age distribution of positive cases in male and female cases tested for each zones shows a significantly more (p<0.05) more positivity in male patients, as shown in *supplementary Figure S2*. Similarly, Male to female ratio was found to be in the same range for all the zones. (Figure 6).

**Figure 5:**
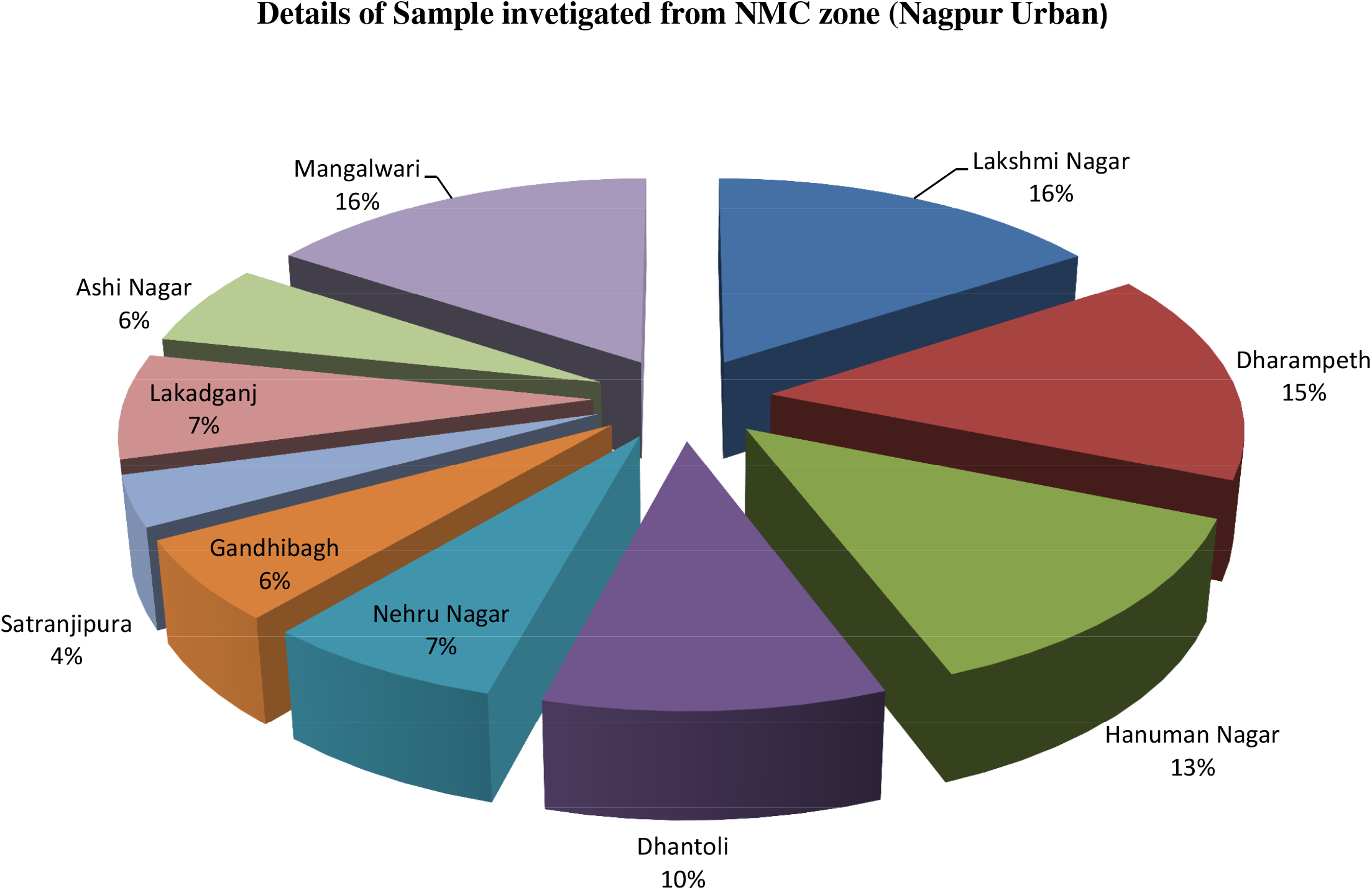
Overall zonal distribution of COVID cases tested.

**Figure 6:**
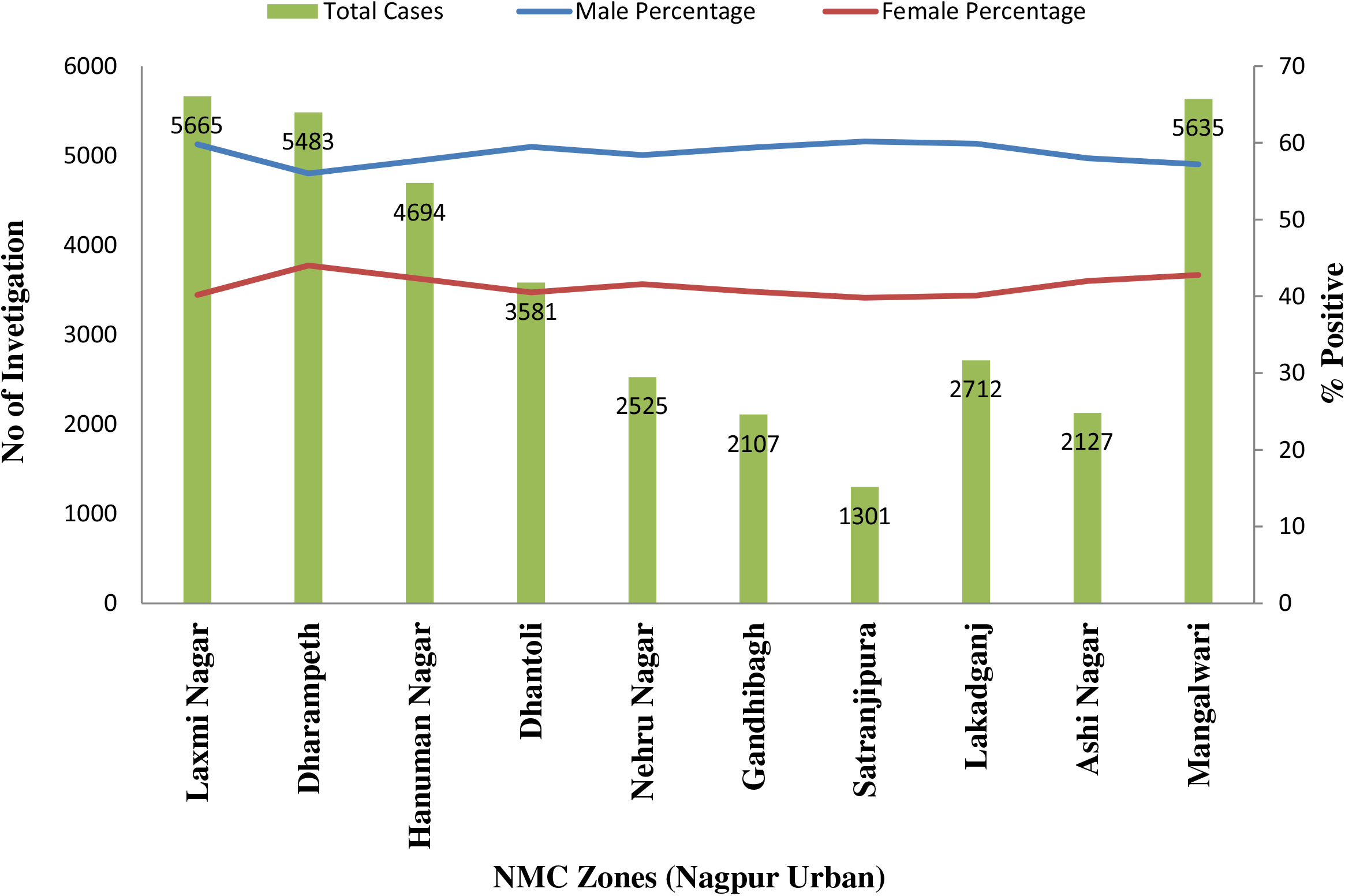
Gender Distribution in different zones of Nagpur

Figure 7 indicates the percentage of male positive cases, female positive cases and total positivity observed among the cases tested in each area. The samples tested in the Satranjipura area showed the highest positivity with 44.43% (p<0.05 vs rest of zone), followed by the Gandhibagh, Ashi Nagar and Lakadganj areas with 42% (p<0.05 vs rest of the zone), 41.09% and 40.63% respectively. Samples received from these zones were less as the samples were sent to other nearby COVID-19 testing centers while positivity was high as these zones were major COVID-19 hotspots in the city and were marked under red zone. Among the cases tested more cases were from the zones like Dharampeth, Lakshmi Nagar, Mangalwari and Dhantoli as Dhruv Lab is located in Dharampeth zone, so more samples were received from this and surrounding zones.

**Figure 7:**
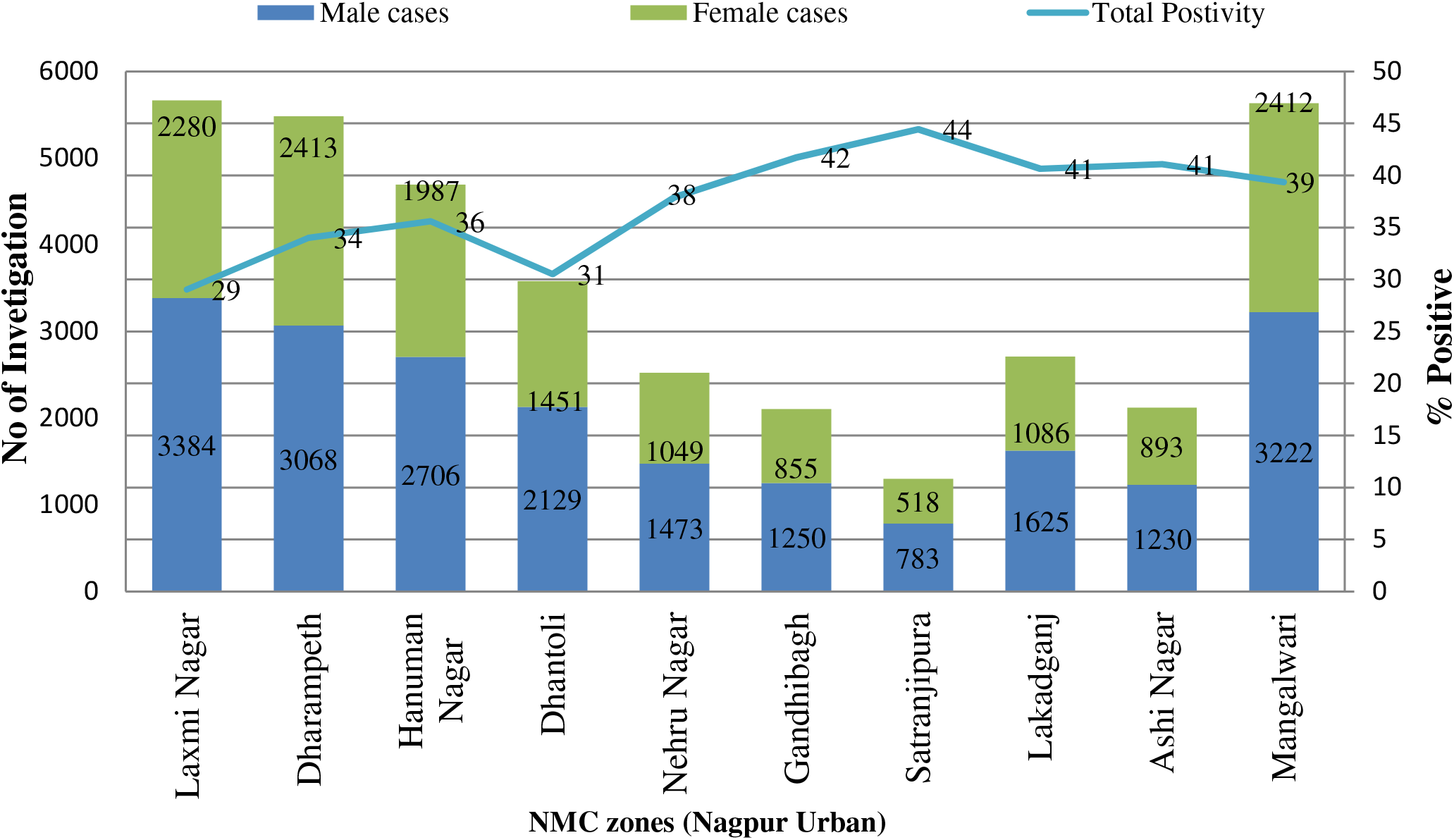
Zone-wise distribution of test results among male and female population tested.

The RT PCR performed was of a qualitative type, so the Ct value did not have any prognostic significance. Ct value also depends on timing of sample collection and quality of sample. However lower Ct value indicates higher viral load. Mean Ct value in male and female cases tested for each zone was calculated to determine case severity and viral load in individual zones as shown in **Figure S3**. It was similar in males and females cases for all the zones and was in the range 22-24. Likewise zonal distribution of mean Ct value among different age groups was also in similar range in all the zones **Figure S4**.

## Discussion

All major countries have experienced COVID-19 as a result of its high transmission rate. The current study shows the COVID-19 RT PCT testing done at a private lab and tracing these cases using the address and pin code details provided while submitting the sample for testing and distribution of these cases for 10 zones of Nagpur. Overall, 34% of positivity was observed from this screening centre over the aforementioned period. The majority of samples were received from the location in the vicinity of the screening facility. Male cases were higher among the total samples received for testing (51%) and positive cases (62%). This is similar to numerous reported studies that show that men are more likely to experience serious symptoms. This is due to the fact that the male population is more exposed compared to the female population [16-18]. The mortality rate for males is also higher than for females [18]. In this study, the RT PCR mean Ct was in the range of 23 in both male and female cases, with little difference. The incidence of COVID-19 is growing at an unprecedented rate across all age groups. In this study, the 21-60 years age group represented 75% of the cases tested and positivity was also high amongst them. In addition, people over the age of 50 with an illness are at higher risk because of low immunity [19, 20].

To detect COVID-19 infection in population RT-PCR testing is the gold standard technique as the specificity is high and sensitivity is moderate compared to other antigen and antibody based assays that is rapid to implement, less expensive compared to other test, easier to perform and do not need much laboratory training, however to confirm diagnosis of COVID-19 infection RT-PCR testing is must even if test result for rapid antigen test is negative [21-23]. Along with these false negative outcomes, COVID-19 screening is a major concern. Although sensitivity and specificity is high for molecular testing, few studies show a false positive result. This depends on the number of factors like high sample processing number, defective kits, instrument fluctuation and contaminated viral medium [24, 25]. Recent studies have also shown that genetic variation may be linked to false positive results higher than expected [26].

Using RT-PCR technique it is possible to detect viable viral particles and determine the viral load in population tested. In order to increase COVID-19 testing, private labs with available infrastructure have been approved by the Government of India and ICMR. In Nagpur, Dhruv Laboratory was the first laboratory to obtain approval for testing, which was helpful for precautionary strategies to control the spread of infection [27]. Collaboration of government and private institutions is a key factor in this crisis situation. Allowing multiple private institutions to conduct testing will increase the rate of daily testing, which is useful for taking action by authorities. In order to increasing COVID testing, test rates were revised by government form may to December 2020 to reduce costing for each samples processed. This includes cost of all the consumables like extraction kit, PCR kit, viral transport medium (VTM), PPE kits, sample transportation etc. Samples were received from various sites such as hospitals, Sample collection centers, quarantine centers and from patient’s residence. Initially, COVID-19 RT-PCR testing was priced at 4,800 per sample. Rates revision are shown in supplementary table (supplementary table 1) which were revised multiple times by the government: in the month of June 2020 and the cost was 2,500/- for each sample, in the month of August 2020 the cost was 2,200/-, in the month of September 2020 the cost was 1,200/-, in the month of October 2020 the cost was 980/-, presently the testing rates are 700/- for each samples [28, 29].

Samples were received from all parts of Nagpur and using the details provided contact search was carried out. Certain parts were major hotspots in the city and the study results matches with the overall trend for Nagpur urban region. This includes areas such as Satranjipura, Gnadhibagh, Mangalwari, Itwari, Mahal, Mominpura etc [30]. These areas are among the densely populated region and as a result transmission was rapid. To contain the spread of infection strict measures such as complete lockdown in city, blocking area with positive cases, travel restriction, mandatory wearing mask, maintaining six feet distancing and night curfews were introduced. [30,31]. This greatly helped to stop the spread of the infection, but due to the economic decline, unlocking has been introduced by Nagpur’s governing authority into areas where there are no cases of COVID-19. This has caused a sharp increase in positive cases and mortality. There was a rise in COVID-19 cases form month of September 2020 [32, 33]. Many cases were asymptomatic as they didn’t show any symptoms related to COVID-19 but the test results were positive. With again surge in the cases there is much need of increasing testing to better understand the spreading dynamics. Vaccination campaigns should begin with hotspot areas, hospitals, test centers and people who are more in contact with daily work.

## Conclusion

The result of present epidemiology study highlights important data with respect to regions of endemicity within Nagpur city zones. The present data has high public health importance and will be useful for local civic bodies and other community stake holders to undertake appropriate control measures in future epidemic waves of Covid 19. Interestingly, the Government’s reduction in testing rates has been helpful in increasing testing per day. The authorization of private laboratories has also increased testing

## Data Availability

NA

## Acknowledgement

None

---

**Table 1:**
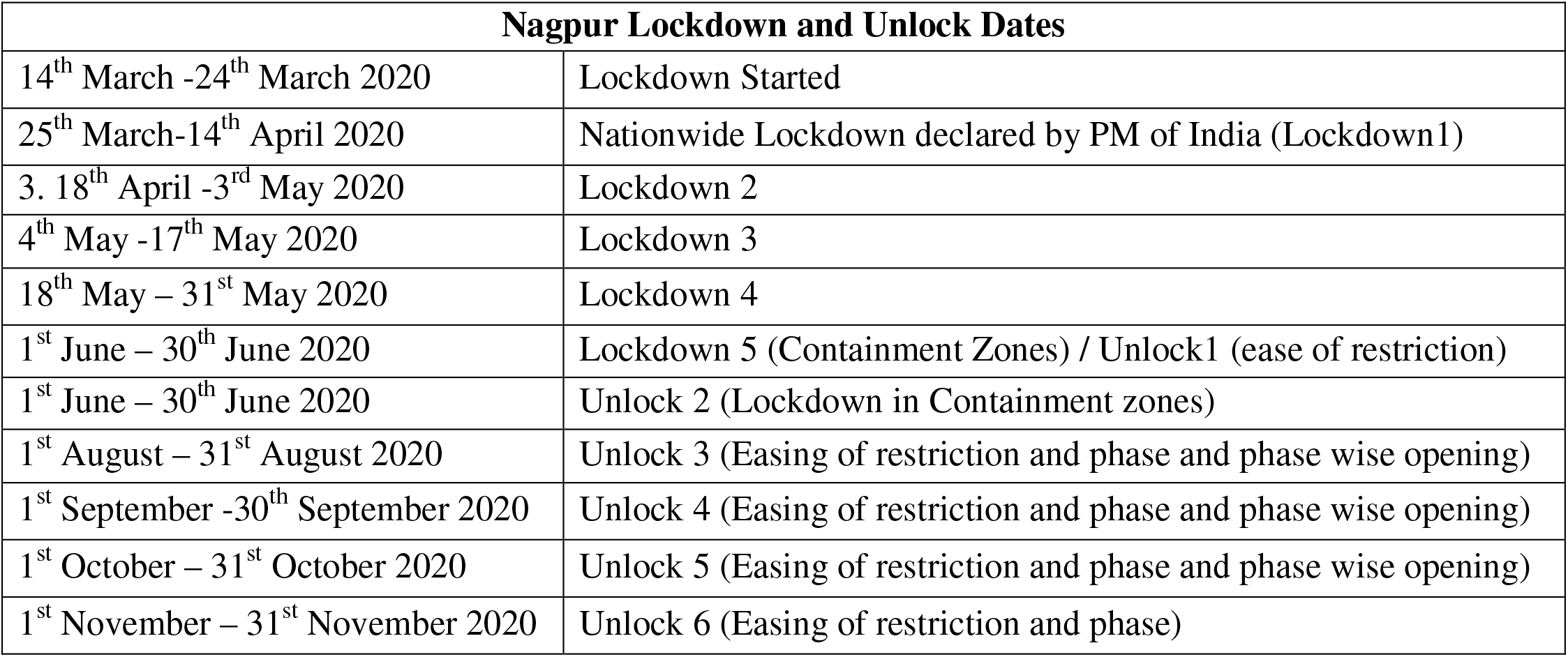
Nagpur Lockdown and Unlock dates for COVID-19

## Supplementary Figures and tables

**Supplementary Table 1:** COVID testing cost revision from May 2020 to December 2020

| Sr. No . | RTPCR test | Beginning (May 2020) | State Govt. Order dated on 13<sup>th</sup> June 2020 | State Govt. Order dated on 30<sup>th</sup> August 2020 | State Govt. Order dated on 9<sup>th</sup> September 2020 | State Govt. Order dated on 10<sup>th</sup> October 2020 | State Govt. Order dated on 14<sup>th</sup> December 2020 |
| --- | --- | --- | --- | --- | --- | --- | --- |
| 1 | Pick of Sample from collection site, transport and reporting of sample (All necessary consumable like VTM, PPE etc. For collection will be borne by sample provider) | 4800/- | 2500/- | 2200/- | 1200/- | 980/- | 700/- |
| 2 | Collection of samples from drive through sample collection kiosks, sample collection drives, covid collection centres, hospital clinic, quarantine centre of lab., etc. (All necessary consumable like VTM, PPE etc. For collection will be borne by laboratory) | 4800/- | 2800/- | 2800/- | 1600/- | 1400/- | 850/- |
| 3 | Collection patient's samples from residence, transport of samples, testing and reporting of sample (All necessary consumable like VTM, PPE etc. For collection will be borne by laboratory) | 4800/- | 2800/- | 2800/- | 2000/- | 1800/- | 980/- |

**Figure S1:**
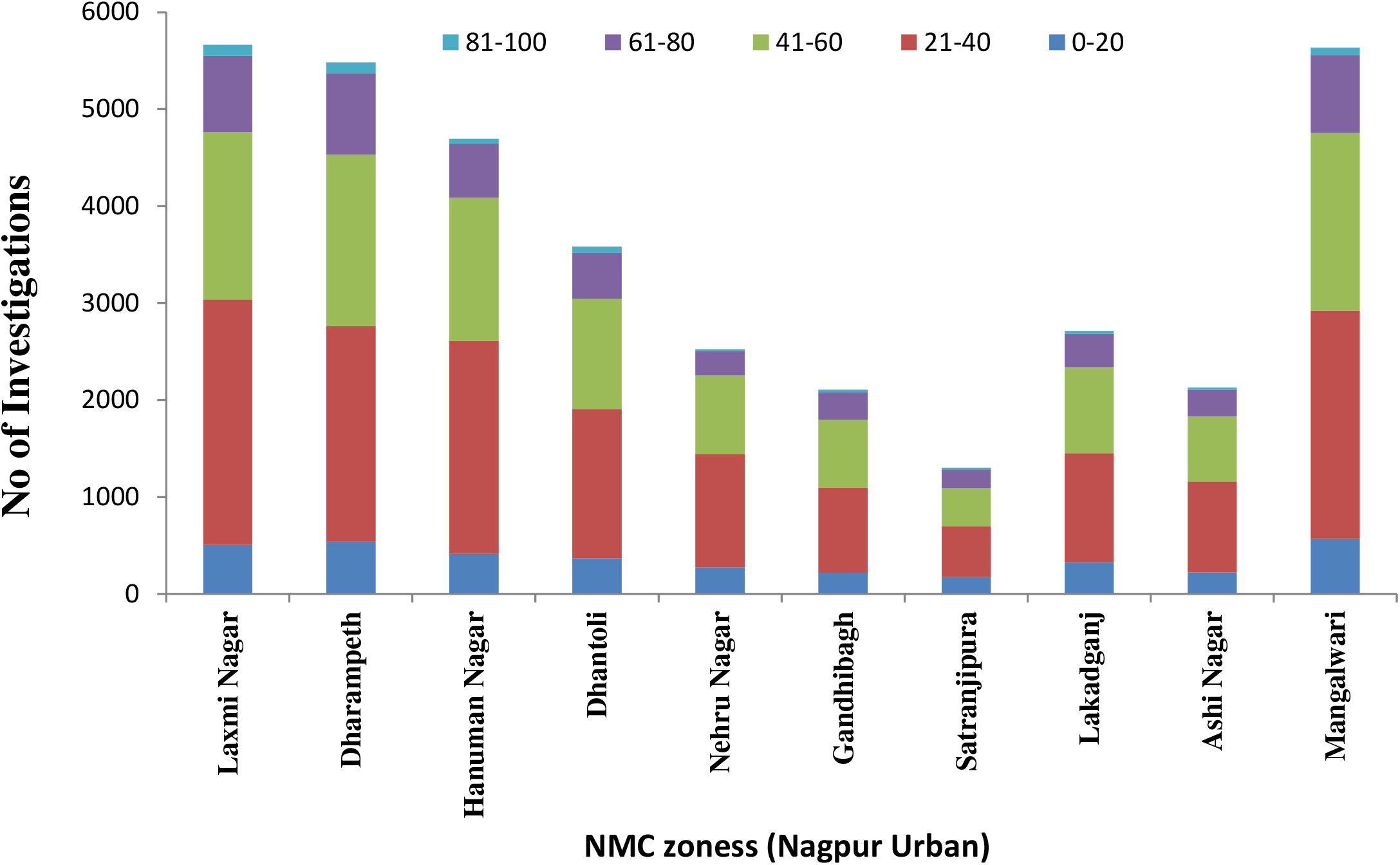
Distribution of cases in different age groups for each zones

**Figure S2:**
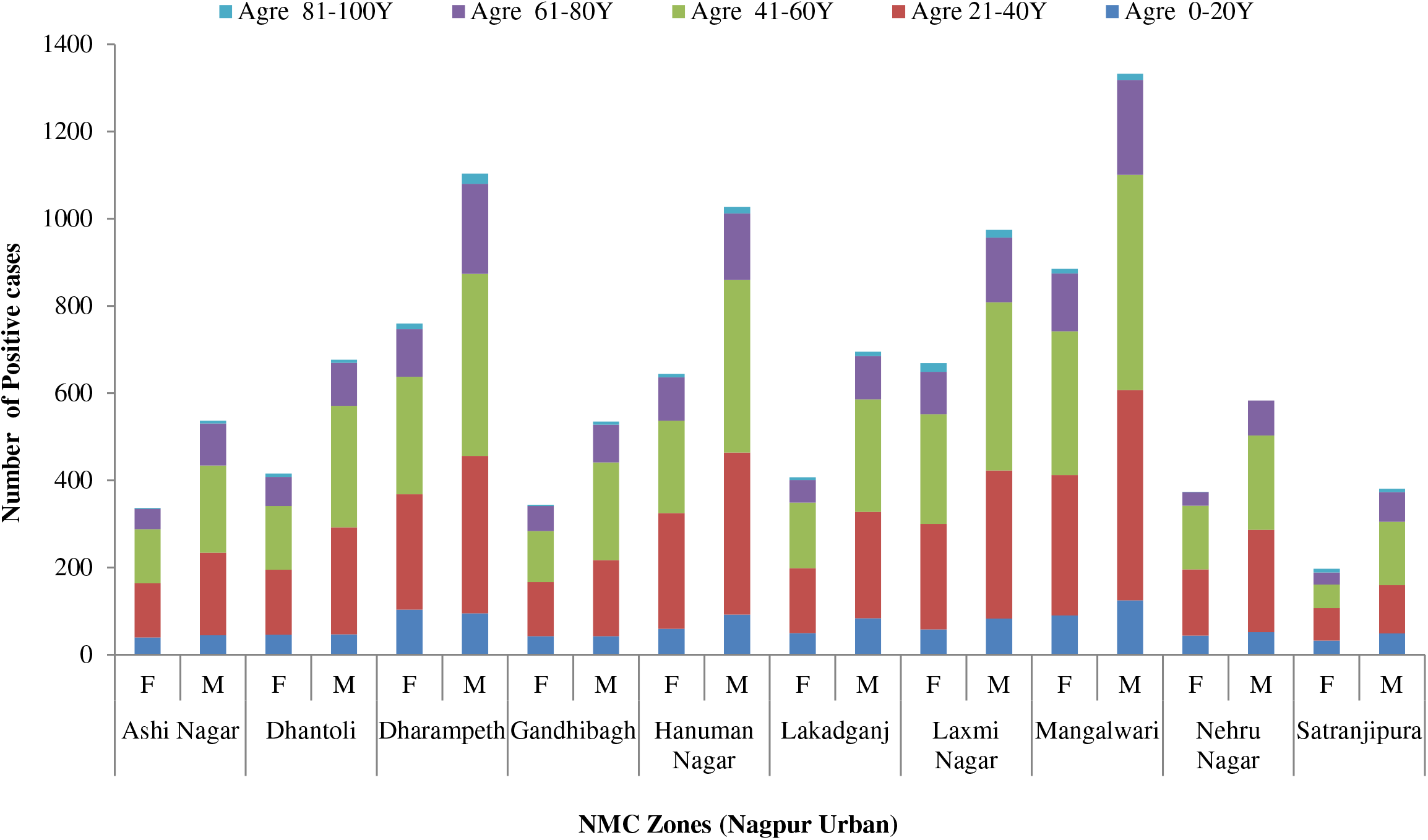
Age distribution of positive cases in male and female cases tested for each zones.

**Figure S3:**
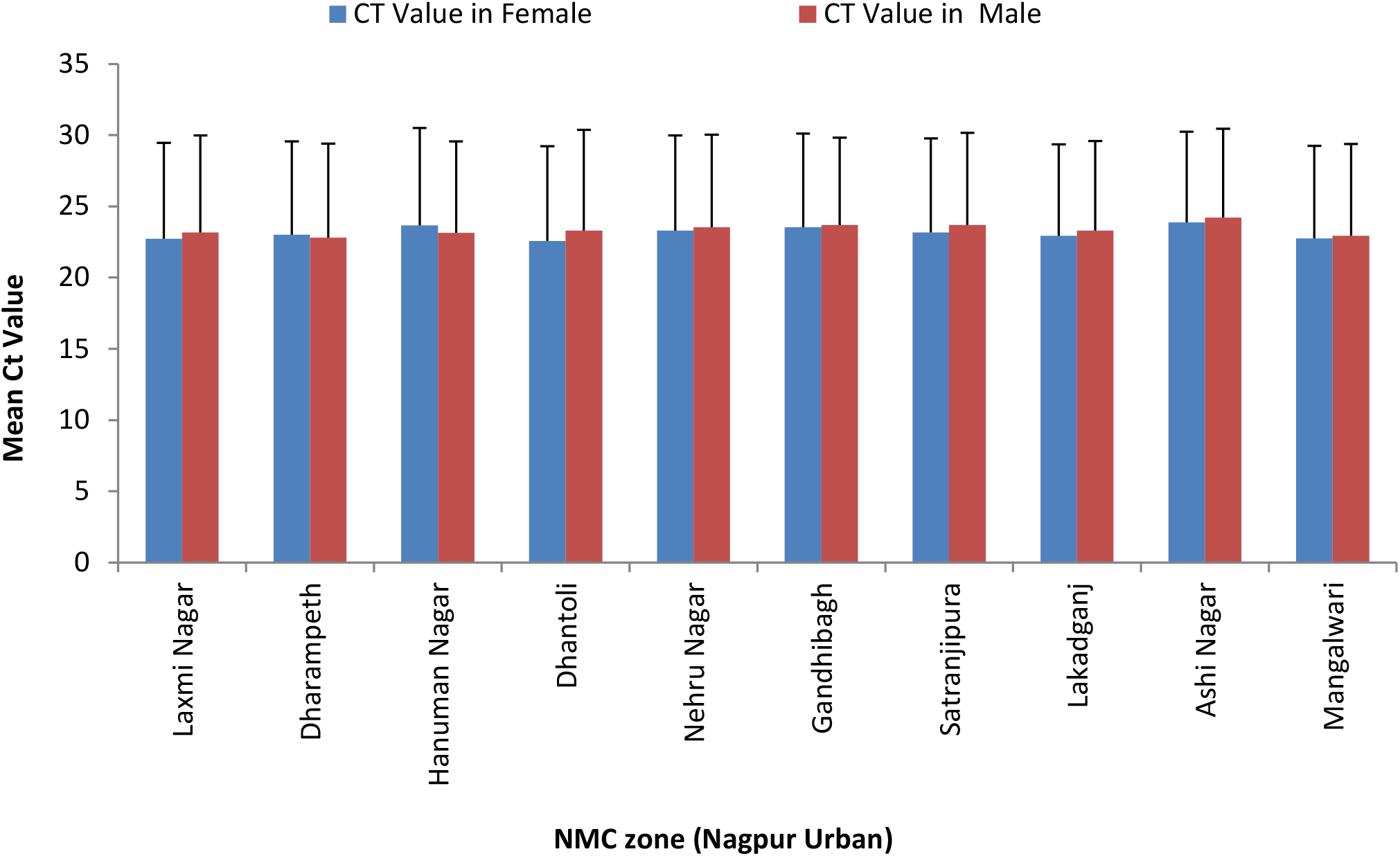
Distribution of Mean Ct Value of positive cases in male and female for each zone

**Figure S4:**
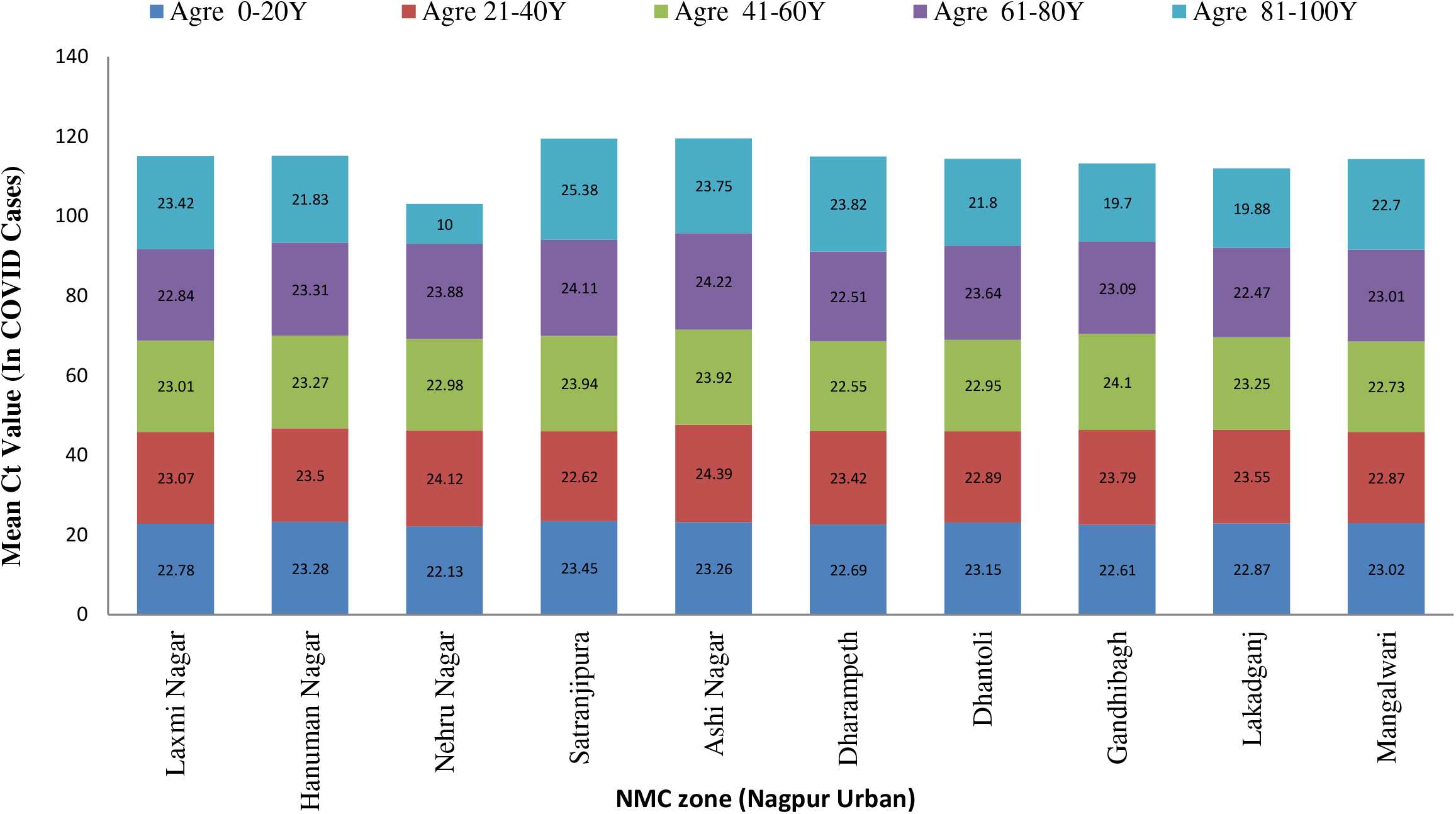
Zonal distribution of mean Ct value in positive cases among different age groups

## References

1. Huang C, Wang Y, Li X, Ren L, et al (2020). Clinical features of patients infected with 2019 novel coronavirus in Wuhan, China. Lancet. 2020 Feb 15;395(10223):497–506.

2. Rothan HA, Byrareddy SN. The epidemiology and pathogenesis of coronavirus disease (COVID-19) outbreak. J Autoimmun. 2020 May; 109:102433.

3. COVID-19 Worldometer update. Available online: www.worldometers.info/coronavirus/ (Cited on 6^th^ April 2020)

4. Andrews, M A et al. “First confirmed case of COVID-19 infection in India: A case report.” The Indian journal of medical research vol. 151,5 (2020): 490-492. doi:10.4103/ijmr.IJMR_2131_20

5. Kodge, B. G.. “A review on current status of COVID19 cases in Maharashtra state of India using GIS: a case study.” Spatial Information Research, 1–7. 27 Jul. 2020,

6. COVID cases in India and state wise representation. (Cited on: 22^nd^ Feb 2021) Available from URL: https://www.covid19india.org/

7. Epidemiology and its role in COVID-19. Available online: https://coe.uni.edu/epidemiology-and-its-role-covid-19 (Cited on: 7th April 2020)

8. Muhammad Shoaib Akhtar. Role of epidemiological studies in disease prevention. The International Journal of Frontier Sciences. 2019 Jan

9. Deshkar S. (2019) Resilience Perspective for Planning Urban Water Infrastructures: A Case of Nagpur City. In: Ray B., Shaw R. (eds) Urban Drought. Disaster Risk Reduction (Methods, Approaches and Practices). Springer, Singapore.

10. Chakraborty P. 10 Zones Restructured based on 38 Wards. The Times of India. Available online:https://timesofindia.indiatimes.com/city/nagpur/10-zones-restructured-based-on-38wards/articleshow/57741271.cms#:~{}:text=Accordingly%2C%20Laxmi%20Nagar%20zone%20will,02%2C%2003%2C%2006%20and%2007 (Cited on: 22 Feb 2021).

11. COVID-19 Monitoring Dashboard by Public Health Department, Government of Maharashtra. Available online: https://arogya.maharashtra.gov.in/ (Cited on: 22^nd^ Feb 2021).

12. Masthi R, Jahan A, Bharathi D, Abhilash P, Kaniyarakkal V, Tv S, Gowda G, Ts R, Goud R, Rao S, Hegde A. Postcode based participatory disease surveillance systems : a comparison with traditional risk-based surveillance and its application in the COVID-19 pandemic. JMIR Public Health Surveill. 2021 Jan 19.

14. Wang Y, Wang Y, Chen Y, Qin Q, Unique epidemiological and clinical features of the emerging 2019 novel coronavirus pneumonia (COVID-19) implicate special control measures. J Med Virol. 2020 Jun; 92(6):568–576

15. Chatterjee P, Anand T, Singh KJ, et al. Healthcare workers & SARS-CoV-2 infection in India: A case-control investigation in the time of COVID-19. Indian J Med Res. 2020 May;151(5):459–467.

16. Li Q, Guan X, Wu P, Wang X, Zhou L et al. Early Transmission Dynamics in Wuhan, China, of Novel Coronavirus-Infected Pneumonia. N Engl J Med. 2020 Mar 26; 382(13):1199-1207.

16. Maleki Dana P, Sadoughi F, Hallajzadeh J, et al. An Insight into the Sex Differences in COVID-19 Patients: What are the Possible Causes?. Prehosp Disaster Med. 2020;35(4):438-441. doi:10.1017/S1049023X20000837

17. Penna C, Mercurio V, Tocchetti CG, Pagliaro P. Sex-related differences in COVID-19 lethality. Br J Pharmacol. 2020 Oct;177(19):4375-4385. doi: 10.1111/bph.15207. Epub 2020 Aug 5. PMID: 32698249; PMCID: PMC7405496

18. Newman CB. Mortality in COVID-19: Further Evidence for a Sex-Based Difference in the OpenSAFELY Study. J Womens Health (Larchmt). 2021 Jan;30 (1):61-63Epub 2020 Dec 8. Erratum in: J Womens Health (Larchmt). 2021 Mar;30(3):452. PMID: 33297829.

19. Chen Y, Klein SL, Garibaldi BT et al. Aging in COVID-19: Vulnerability, immunity and intervention. Ageing Res Rev. 2021 Jan;65:101205

20. O’Driscoll M, Ribeiro Dos Santos G, Wang L et al. Age-specific mortality and immunity patterns of SARS-CoV-2. Nature. 2021 Feb;590(7844):140–145.

22. Dramé M, Tabue Teguo M, Proye E, Hequet F et al. Should RT-PCR be considered a gold standard in the diagnosis of COVID-19? J Med Virol. 2020 Nov;92(11):2312–2313.

23. Oliveira BA, Oliveira LC, Sabino EC, Okay TS. SARS-CoV-2 and the COVID-19 disease: a mini review on diagnostic methods. Rev Inst Med Trop Sao Paulo. 2020 Jun 29;62:e44. PMID: 32609256; PMCID: PMC7325591.

24. Kanji JN, Zelyas N, MacDonald C et al. False negative rate of COVID-19 PCR testing: a discordant testing analysis. Virol J. 2021 Jan 9;18(1):13. PMID: 33422083; PMCID: PMC7794619.

25. Woloshin S, Patel N, Kesselheim AS. False Negative Tests for SARS-CoV-2 Infection - Challenges and Implications. N Engl J Med. 2020 Aug 6;383(6):e38. Epub 2020 Jun 5. PMID: 32502334.

26. Genetic Variants of SARS-CoV-2 May Lead to False Negative Results with Molecular Tests for Detection of SARS-CoV-2 - Letter to Clinical Laboratory Staff and Health Care Providers. Available online: https://www.fda.gov/medical-devices/letters-health-care-providers/genetic-variants-sars-cov-2-may-lead-false-negative-results-molecular-tests-detection-sars-cov-2 (Cited on: 7th April 2020)

27. City’s ‘Dhruv’, first private lab in region authorised to test Covid-19 samples. Available online: https://www.thehitavada.com/Encyc/2020/5/3/City-s-Dhruv-first-private-lab-in-region-authorised-to-test-Covid-19-samples.html (Cited on: 19th Feb 2021)

28. Debroy S. COVID-19 Maharashtra slashes testing rates for RT-PCR,rapid antigen tests Available online: https://timesofindia.indiatimes.com/city/mumbai/covid-19-maharashtra-slashes-testing-rates-for-rt-pcr-rapid-antigen-tests/articleshow/79744709.cms (Cited on: 19th Feb 2021)

29. Maharashtra govt reduces COVID RT-PCR test cost from Rs 980 to Rs 700. Available online: https://www.timesnownews.com/india/article/maharashtra-govt-reduces-covid-rt-pcr-test-cost-from-rs-980-to-rs/695019 (Cited on: 19th Feb 2021)

30. Now, 8 areas in districts have 10 or more COVID-19 cases (Published online: 13^th^ June 2020). Available online: https://timesofindia.indiatimes.com/city/nagpur/now-8-areas-in-district-have-10-or-more-covid-19-cases/articleshow/76348705.cms (Cited on: 5th April 2021

31. Four Maharashtra cities including Pune, Mumbai, Nagpur in lockdown due to COVID-19. Available online: https://www.indiatvnews.com/news/india/four-maharashtra-cities-including-pune-mumbai-nagpur-lockdown-covid-19-599919 (Published online: 20^th^ March 2020) (Cited on: 5^th^ April 2021)

31. Saha, Jay et al. “Lockdown for COVID-19 and its impact on community mobility in India: An analysis of the COVID-19 Community Mobility Reports, 2020.” Children and youth services review vol. 116 (2020): 105160. doi:10.1016/j.childyouth.2020.105160

32. Mission Begin Again” Maharashtra set for unlock 1.0. Available online: https://thelivenagpur.com/2020/05/31/mission-begin-again-maharashtra-set-for-unlock-1-0/ (Cited on 5^th^ April 2021)

33. Nagpur reports highest one-day spike of 2,343 cases. (Published online: 13^th^ September 2020) Available online:outlookindia.com/newsscroll/covid19-nagpur-reports-highest-oneday-spike-of-2343-cases/1935047 (Cited on 5^th^ April 202

